# Effects of social distancing on the spreading of COVID-19 inferred from mobile phone data

**DOI:** 10.1101/2020.06.25.20140434

**Authors:** Hamid Khataee, Istvan Scheuring, Andras Czirok, Zoltan Neufeld

**Affiliations:** School of Mathematics and Physics, The University of Queensland, St. Lucia, Brisbane, QLD 4072, Australia; Evolutionary Systems Research Group, Centre for Ecological Research, Hungarian Academy of Sciences, Tihany, Hungary; MTA-ELTE Theoretical Biology and Evolutionary Ecology Research Group, Hungarian Academy of Sciences, Budapest, Hungary; Department of Biological Physics, Eotvos University, Budapest, 1053, Hungary; Department of Anatomy and Cell Biology, University of Kansas Medical Center, Kansas City, KS 66160, USA

## Abstract

A better understanding of how the COVID-19 epidemic responds to social distancing efforts is required for the control of future outbreaks and to calibrate partial lock-downs. We present quantitative relationships between key parameters characterizing the COVID-19 epidemiology and social distancing efforts of nine selected European countries. Epidemiological parameters were extracted from the number of daily deaths data, while mitigation efforts are estimated from mobile phone tracking data. The decrease of the basic reproductive number (*R*_0_) as well as the duration of the initial exponential expansion phase of the epidemic strongly correlates with the magnitude of mobility reduction. Utilizing these relationships we decipher the relative impact of the timing and the extent of social distancing on the total death burden of the epidemic.

## 1 Introduction

The COVID-19 epidemic started in late 2019 and within a few months it spread around the World infecting 9 million people, out of which half a million succumbed to the disease. As of June 2020, the transmission of the disease is still progressing in many countries, especially in the American continent. While there have been big regional differences in the extent of the epidemic, in most countries of Europe and Asia the initial exponential growth has gradually transitioned into a decaying phase. An epidemic outbreak can recede either due to reduction of the transmission probability, or due to a gradual build up of immunity within the population. According to currently available immunological data [1, 2] at most locations only a relatively small fraction of the population was infected, typically well below 10%, thus the receding disease mostly reflects changes in social behavior and the associated reduction in disease transmission. Changes in social behavior can include state-mandated control measures as well as voluntary reduction of social interactions. However, especially with the view of potential future outbreaks, it is important to better understand how the timing and extent of social distancing can impact the dynamics of the COVID-19 epidemic.

Previous studies estimated the effect of social distancing on the dynamics of COVID-19 epidemics either by direct data analysis or by modeling methods. A statistical analysis of the number of diagnosed cases, deaths and patients in intensive care units (ICU) in Italy and Spain have indicated that the epidemic started to decrease only after the introduction of strict lock-down action [3]. This was especially visible in Italy, where the final strict social distancing has been reached in a number of consecutive steps. A statistically more comprehensive analysis of hospitalized and ICU patients in France identified a 77% decrease in the growth rate of these numbers after the introduction of the lock-down [4]. The comparison of social distancing efforts in China, South Korea, Italy, France, Iran and USA [5] revealed that the initial doubling time of identified cases was about 2 days which was reduced substantially by the various restrictions introduced in these countries. Epidemic models are widely used to estimate how various intervention strategies affect the transmission rate, however, the quantitative relationship between social interventions and epidemic parameters are hardly known. [6, 7].

In this paper, we quantitatively characterise the time course of the COVID-19 epidemic using daily death data from nine selected European countries [8]. Statistical data on COVID-19-related deaths are considered to be more robust than that of daily cases of new infections. The latter is affected by the number of tests performed as well as by the testing strategy – e.g. its restriction to symptomatic patients – which may be highly variable across countries and often changes during the course of the epidemic. The time course of daily deaths can be considered as a more reliable indirect delayed indicator of daily infections. We thus do not address apparent differences in the case fatality ratio, and restrict our focus to the recorded COVID-19-associated death toll.

Our choice of countries was motivated by the requirements that each of these countries (i) had a relatively large disease-associated death toll (i.e. typically above 10/day and more than 2000 overall) so we can assume that the deterministic component of the epidemic dynamics dominates over random fluctuations. (ii) The selected countries spent a suitably long time in the decaying phase of the epidemic, thus allowing its precise characterisation. Based on these two criteria, we analysed data from the following, socio-economically similar countries, each implementing a somewhat distinct social distancing response: Italy, Spain, France, UK, Germany, Switzerland, Netherlands, Belgium and Sweden.

To characterise social distancing responses we used mobile phone mobility trend data from Apple Inc. [9]. Our aim is to quantitatively determine the characteristic features of the progression of the epidemic, such as initial growth rate, timing of the peak, and final decay rate and investigate how these parameters are determined by the timing and strictness of social distancing measures. We demonstrate that the overall death burden of the epidemic can be well explained by both the timing and extent of social distancing, often organized voluntarily well in advance of the state-mandated lock-down, and present quantitative relationships between changes in epidemiological parameters and mobile phone motility data.

## 2 Characterization of the epidemic

Daily COVID-19 death data, *D*(*t*), is presented in Fig. 1 for the nine countries analysed. We selected the *t* = 0 reference time point as the date when the daily death rate first exceeded 5 deaths. In each country *D*(*t*) indicates the presence of a well defined exponential growth phase, followed by a crossover region and later an exponential decay stage. The initial growth and final decay regions are characterized by fitting the exponential 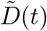 as

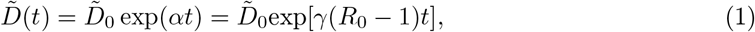

where the fitted functions are distinguished from the actual time series using a tilde, *R*_0_ is the basic reproductive number (i.e. number of new secondary infections caused by a single infected in a fully susceptible population) [10] and *γ* is the inverse average duration of the infectious period. For calculating the basic reproductive number we use *γ* = 0.1 day^−1^. The exact value of *γ* is somewhat uncertain at present [11, 12], however our analysis and results do not rely on the value of this parameter.

**Figure 1:**
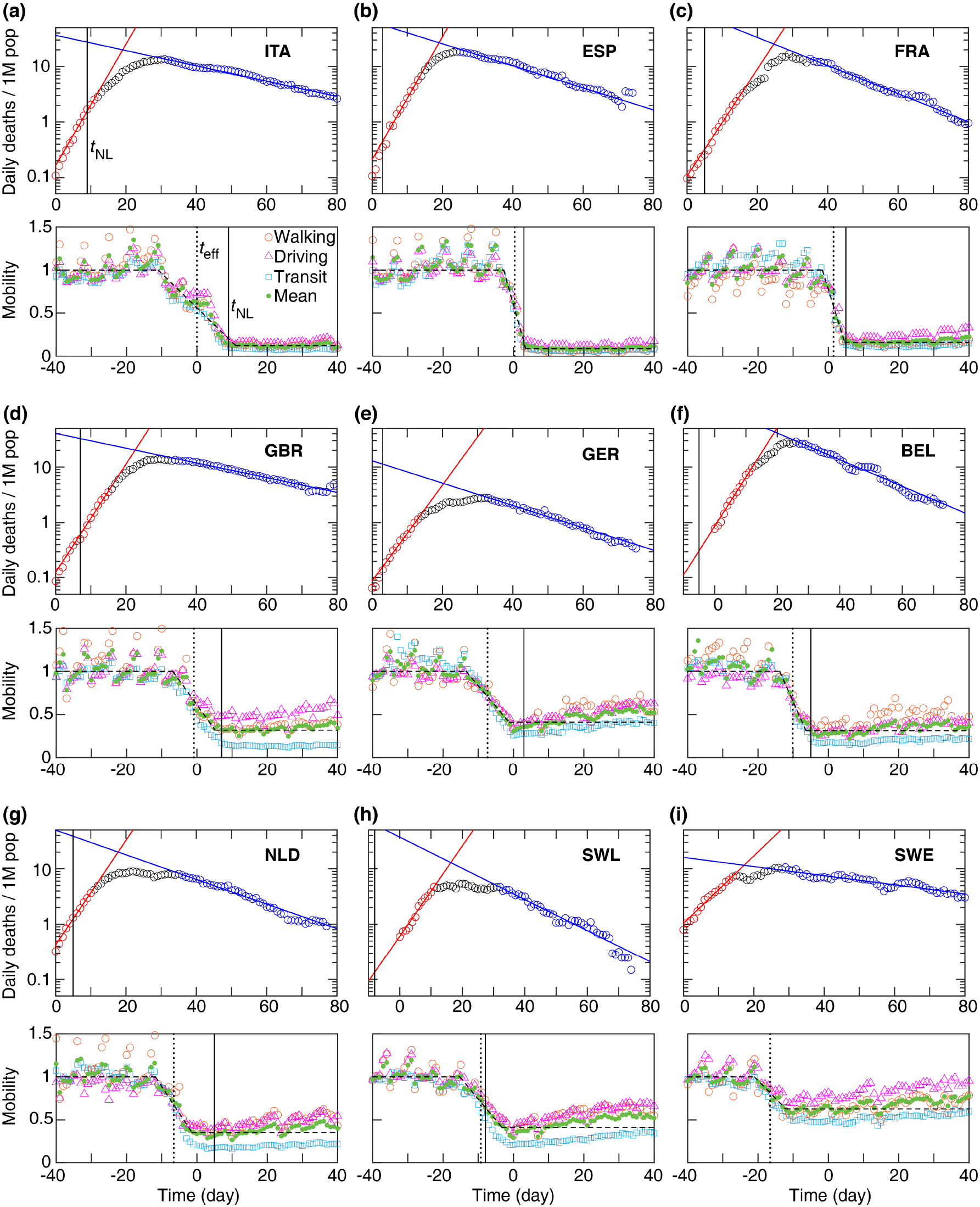
Daily death and mobility data for 9 European countries (a-i). Time 0 corresponds to the day when a country first reported ≥ 5 daily deaths. Top: Growth and decay phases (red and blue lines) were fitted using Equation (1) to the data points highlighted by red and blue, respectively. Vertical line: national lock-down date *t*_NL_. Bottom: Mobile phone tracking data, normalized by the average values before the epidemic (*M*_1_). Quantitative parameters were extracted by a fit (dashed line) to the mean mobility data (green circles, average of walking, driving, and transit data) calculated using Equation (3). Dotted vertical line indicates the effective lock-down date *t*_eff_ calculated using Equation (4). Fit parameters are summarised in Tables S2 and S3.

The values 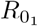] and 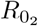, as given in Table 1, thus characterize the growth and decay phases, respectively. The initial growth rate 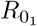 is similar in the selected countries except in Sweden where it is substantially lower. This difference may reflect weaker social mixing, different cultural habits or a somewhat lower population density – an interesting problem outside of the scope of this report. The reproductive number in the decaying phase 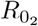 is more variable across the different countries: in particular the decay is significantly slower in Sweden and somewhat slower in the UK.

**Table 1:** Key parameters of the daily COVID-19 death toll and social distancing.

| Country | $R_{01}$ | $R_{02}$ | $t_c$ | $t_{\text{eff}}$ | $t_c - t_{\text{NL}}$ | $t_c - t_{\text{eff}}$ | $\mu$ |
| --- | --- | --- | --- | --- | --- | --- | --- |
| Italy (ITA) | 3.52 | 0.68 | 19.07 | 0.004 | 10.07 | 19.07 | 0.12 |
| Spain (ESP) | 3.54 | 0.54 | 19.03 | 0.38 | 16.03 | 18.65 | 0.09 |
| France (FRA) | 3.22 | 0.42 | 24.66 | 1.47 | 19.66 | 23.19 | 0.16 |
| Great Britain (GBR) | 3.24 | 0.70 | 22.60 | -0.2 | 15.60 | 23.42 | 0.32 |
| Germany (GER) | 3.00 | 0.54 | 20.32 | -7.33 | 17.32 | 27.65 | 0.41 |
| Belgium (BEL) | 3.04 | 0.44 | 19.17 | -10.09 | 24.17 | 29.26 | 0.31 |
| Netherlands (NLD) | 3.16 | 0.48 | 17.77 | -6.59 | 12.77 | 24.36 | 0.35 |
| Switzerland (SWL) | 2.88 | 0.36 | 16.32 | -9.26 | 24.32 | 25.58 | 0.41 |
| Sweden (SWE) | 2.40 | 0.82 | 17.23 | -16.53 | - | 33.76 | 0.63 |

A third parameter characterizes the transition from the growth to decay phases, i.e., the peak of the epidemic in terms of deaths. We define 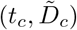 as the intersection point of the two fitted exponential functions. This is a more robust estimator than the actual maximum in daily deaths *D*_*c*_ as at the time of transition daily deaths can exhibit a plateau, hence the value and location of the maximum is sensitive to stochastic fluctuations. The gradual transition from exponential growth to decay in the actual data *D*(*t*) can be due to gradual implementation of social distancing as well as to the case-to-case variability of the time elapsed from infection to death.

The total death toll of the disease can be estimated analytically using the three parameters 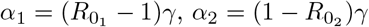 and 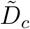 as

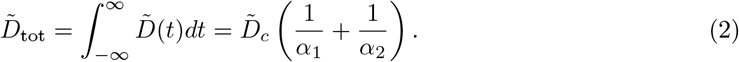

As Fig. 2 demonstrates, despite the substantial variation in death toll among the nine countries, it can be fairly well estimated by the expression (2). Specifically, details of the cross-over region, which do not fit well to the two exponential functions, contribute only around 10% to the overall death toll, while variations in the three epidemiological parameters can change the death burden by an order of magnitude.

**Figure 2:**
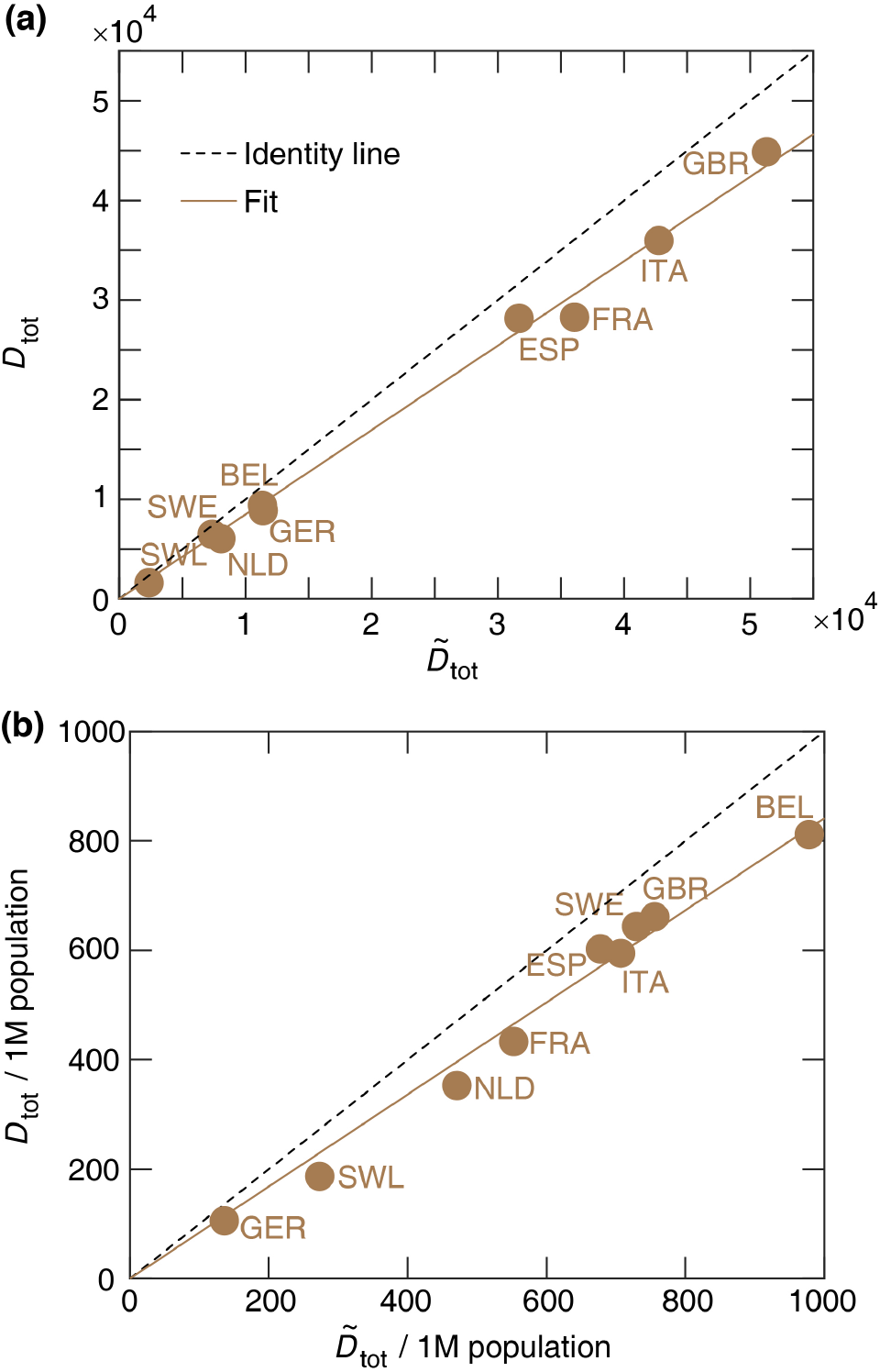
Analytical estimate of the total number of COVID-19 deaths 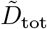 versus *D*_tot_, the actual total death toll (a). (b) The same data are presented as per million (1M) population. The dashed and solid lines represent the identity and a linear regression, respectively. The slope of the linear regression is 0.85 ± 0.01 (a) and 0.84 ± 0.02 (b). Pearson coefficients of determination are (a) *r*^2^ = 0.99 (*p <* 0.001) and (b) *r*^2^ = 0.98 (*p <* 0.001).

## 3 Characterization of social distancing

The timing and strictness of the often voluntary social distancing is quantified from the average mobility data (green circles in Fig. 1), by fitting the following piece-wise linear function:

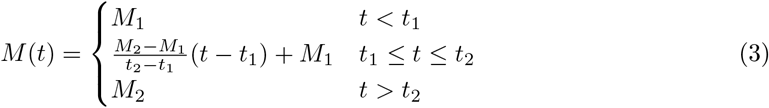

where *M*_*1*_ and *M*_2_ are the average mobility levels before and after social distancing (i.e., before *t*_1_ and after *t*_2_), respectively. To fit Equation (3) to the mobility data, we used the data recorded over 90 days starting from 13-January-2020. In this period the data indicates the mobility levels before and after implementing the social distancing. The walking, driving and transit data are first averaged, then Equation (3) was fitted to the average mobility level. The mobility data with average pre-epidemic value scaled to unity, *M* (*t*)*/M*_1_, is shown in Fig. 1. We characterize the extent of social distancing by the ratio *µ* = *M*_2_*/M*_1_ *<* 1. A strict restriction of social interaction is expected to be reflected as *µ ≈* 0.

Using the fitted parameters *t*_1_ and *t*_2_ obtained from Equation (3), an effective social distancing date (Table 1) is estimated as

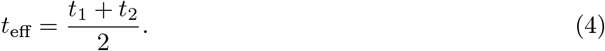

The fitted values of *M*_1_ and *M*_2_ as well as of *t*_eff_ are presented in Fig. 1. It is noteworthy that *t*_eff_ often *preceded* the official national lock-down *t*_NL_.

## 4 The effect of social distancing on epidemic parameters

Next, we investigate how the timing and extent of the social distancing changes the epidemic parameters, and as a consequence, the total death toll. First, we note that surprisingly neither the date of the official national lock-down *t*_NL_ nor the effective date of social distancing predicts well – in itself – the time of the epidemic peak *t*_*c*_ (Fig. 3). As the average time between infection and succumbing to COVID-19 is 18 days [13], a delay of approximately 18 days is expected (dashed line in Fig. 3) between the introduction of social distancing and the change in the trend of the daily death count. Instead, we find that the time to the peak from the official national lock-down *t*_*c*_−*t*_NL_ varies in a range from 10 days in Italy to more than 3 weeks in case of Switzerland, and the time from the change in mobility to the peak, *t*_*c*_−*t*_eff_, ranges from 19 days (Italy and Spain) up to 34 days (Sweden) as shown in Table. 1.

**Figure 3:**
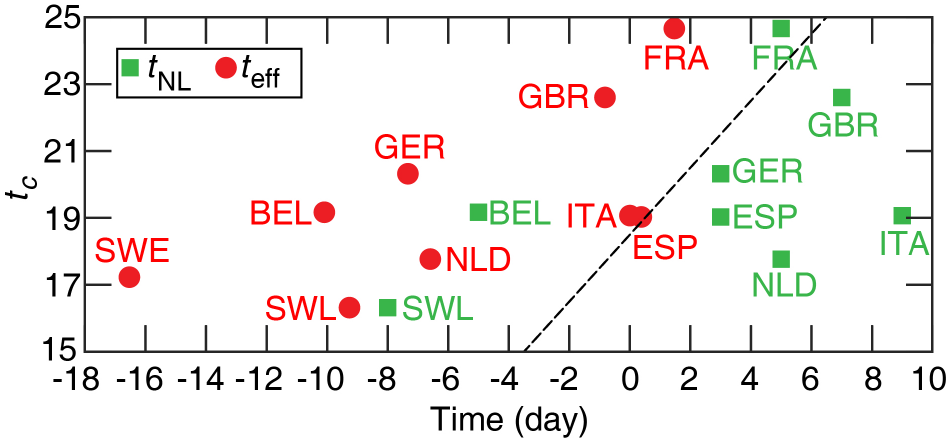
The time of the peak in daily deaths *t*_*c*_ versus the official national lock-down date *t*_NL_ and the effective social distancing date *t*_eff_. The correlation is weak with *r*^2^ = 0.45 (*p* = 0.06) for *t*_eff_, and *r*^2^ = 0.12 (*p* = 0.37) for *t*_NL_. Neither correlations are statistically significant. Dashed line: time (identity line) with a delay of 18.5 days.

Furthermore, neither the time of the peak *t*_*c*_ nor the parameters characterising the time and strength of social distancing *t*_eff_, *t*_NL_, *µ* correlates – as a single parameter – well with the total number of deaths (Fig. 4). For example, Belgium registers a very high per capita death toll despite an early official lock-down, 5 days before the reference time *t* = 0, and even earlier change is observed in mobility data (*t*_eff_ = − 10 days). On the other hand, a moderate social distancing in Germany, indicated by the mobility ratio *µ* = 0.4, led to the lowest death toll within this group of countries. Thus, we decided to investigate more carefully how social distancing affects the three epidemic parameters 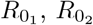 and 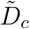 – with the hypothesis that the non-linear, multifactor relationship (2) effectively masks correlations between the death toll and any single control parameter.

**Figure 4:**
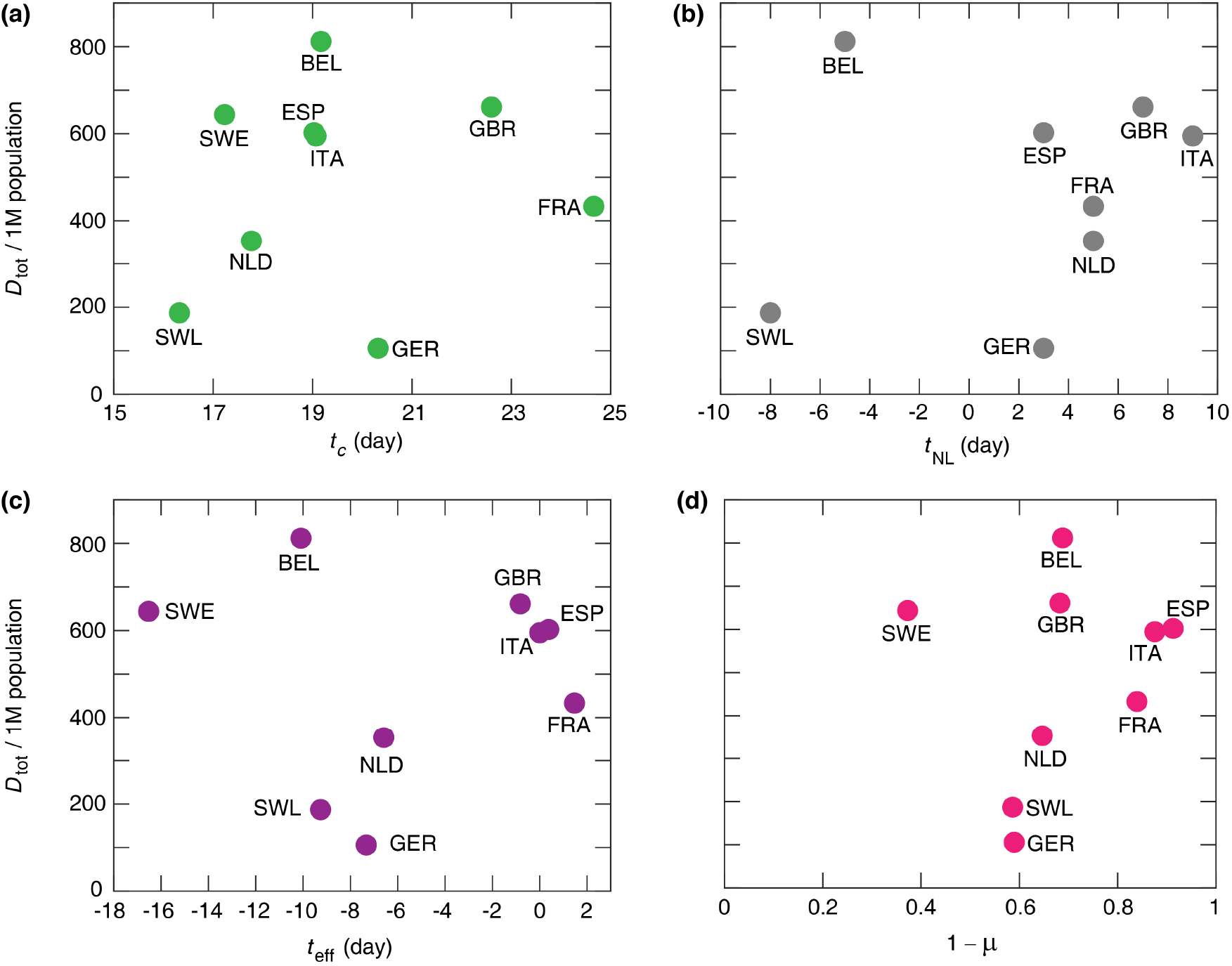
Actual total number of deaths per million (1M) population versus *t*_c_, the peak time of daily deaths (a), *t*_NL_ national lock-down date (b), *t*_eff_ effective lock-down date (c), and 1 − *µ*, the relative mobility drop (d). Pearson correlation coefficients are (a) *r*^2^ = 0.01 (*p* = 0.77), (b) *r*^2^ = 0.06 (*p* = 0.52), (c) *r*^2^ = 0.002 (*p* = 0.92), and (d) *r*^2^ = 0.04 (*p* = 0.62) indicating that there is no statistically significant correlation among these epidemic characteristics.

As Fig. 5 indicates, we found two strong relationships between the epidemiological parameters and measures of social distancing. Figure 5(a) indicates a strong positive correlation between the drop in basic reproductive number,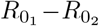 and the restriction of mobility *µ*. The relationship can be well approximated by the quantitative formula:

**Figure 5:**
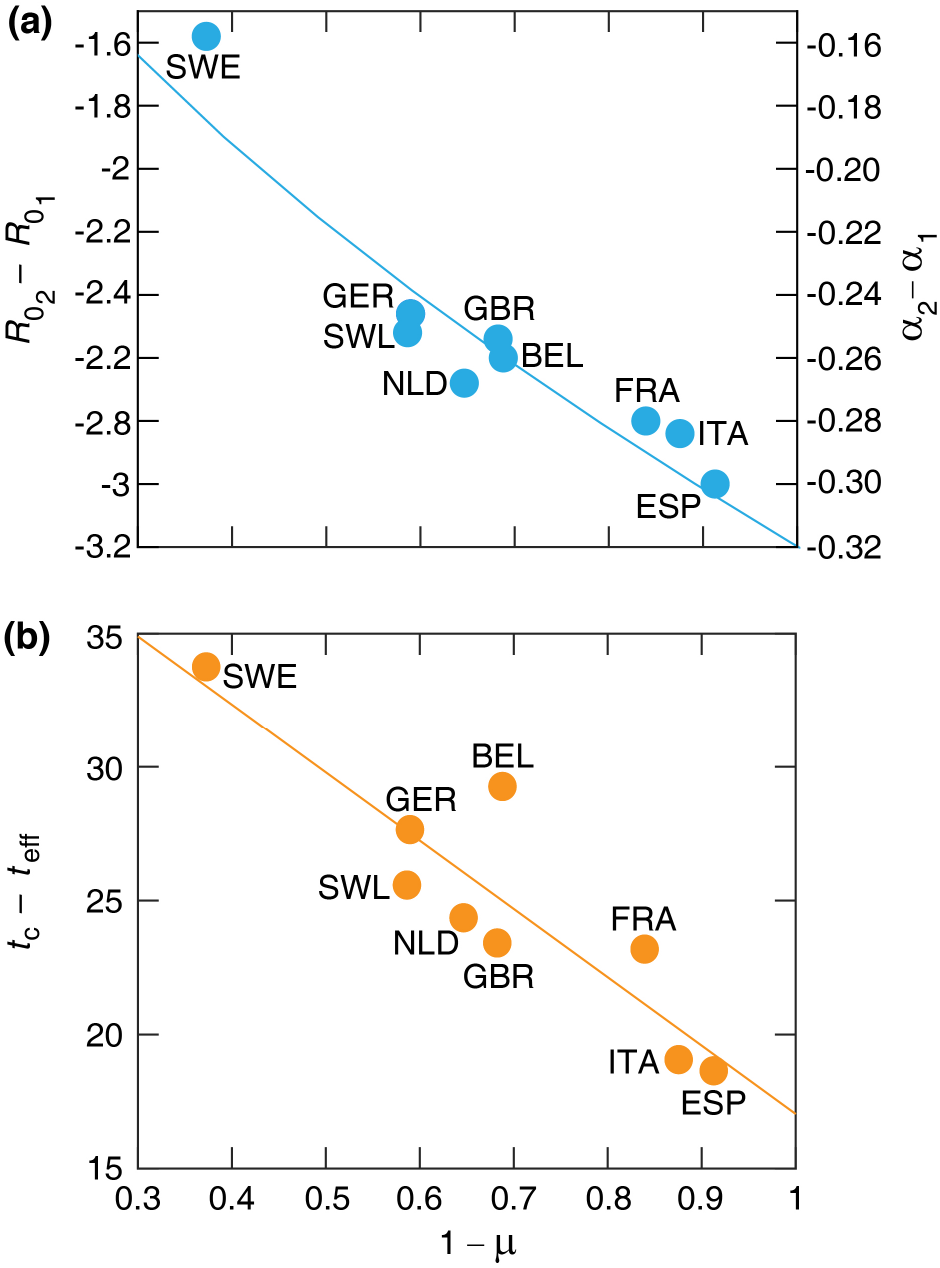
Relationships between key parameters of the COVID-19 epidemics and mobility data. (a) Change of basic reproductive number 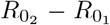 (left axis) and *α*_2_ − *α*_1_ (right axis) versus the motility drop 1 − *µ*. Solid line indicates a power-law fit −*β*(1 − *µ*)^*ρ*^, where *β* = 3.18 ± 0.11 and *ρ* = 0.56 ± 0.09. *r*_2_ = 0.85, (*p* = 0.0005). (b) Elapsed time between the epidemic peak and the effective lock-down, *t*_*c*_ − *t*_eff_, versus the motility drop 1 − *µ*. Solid line indicates the linear fit Equation (6), where *τ*_0_ = 17.04 ± 1.62 and *η* = 1.50 ± 0.40.

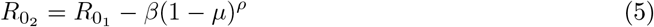

with *ρ* = 0.56 ± 0.09 and *β* = 3.18 ± 0.11. Furthermore, the time elapsed between the peak and the social distancing, *t*_*c*_ − *t*_eff_, correlates negatively with the severity of the mobility restrictions (Fig. 5(b)) as

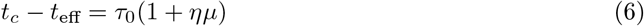

where *τ*_0_ = 17.04±1.62 days is comparable with the mean value of patient survival in fatal COVID-19 disease [13], and *η* = 1.50 ± 0.40 is the factor characterizing the lengthening of the delay for less severe reduction of mobility. Thus, the peak follows strict lock-downs (small *µ* as in Italy and Spain) by around 18 days. For less restrictive social distancing (*µ ≈* 0.6 as in Sweden), however, the peak can be delayed by as much as 5 weeks. This delay is of crucial importance, as the peak of the daily death toll 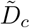 is determined as

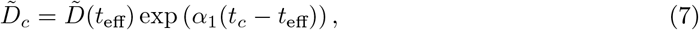

where 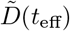 is a natural measure of how early the social distancing took place relative to the dynamics of the epidemics. As 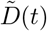 is the exponential fit Equation (1) instead of the actual daily death count *D*(*t*), unfortunately it is difficult to know in real-time. The country-specific values of 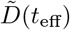 are listed in Table 2.

**Table 2:**
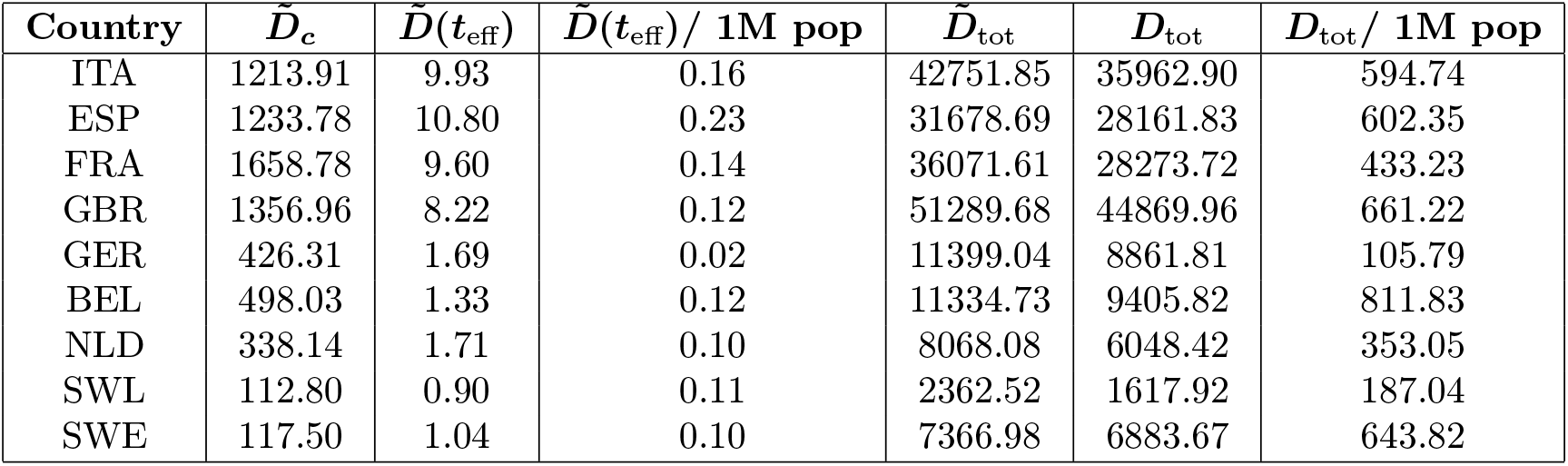
Parameters characterising the death toll of the epidemic.

| Country | $\tilde{D}_c$ | $\tilde{D}(t_{\text{eff}})$ | $\tilde{D}(t_{\text{eff}})/1\text{M pop}$ | $\tilde{D}_{\text{tot}}$ | $D_{\text{tot}}$ | $D_{\text{tot}}/1\text{M pop}$ |
| --- | --- | --- | --- | --- | --- | --- |
| ITA | 1213.91 | 9.93 | 0.16 | 42751.85 | 35962.90 | 594.74 |
| ESP | 1233.78 | 10.80 | 0.23 | 31678.69 | 28161.83 | 602.35 |
| FRA | 1658.78 | 9.60 | 0.14 | 36071.61 | 28273.72 | 433.23 |
| GBR | 1356.96 | 8.22 | 0.12 | 51289.68 | 44869.96 | 661.22 |
| GER | 426.31 | 1.69 | 0.02 | 11399.04 | 8861.81 | 105.79 |
| BEL | 498.03 | 1.33 | 0.12 | 11334.73 | 9405.82 | 811.83 |
| NLD | 338.14 | 1.71 | 0.10 | 8068.08 | 6048.42 | 353.05 |
| SWL | 112.80 | 0.90 | 0.11 | 2362.52 | 1617.92 | 187.04 |
| SWE | 117.50 | 1.04 | 0.10 | 7366.98 | 6883.67 | 643.82 |

Our analysis thus indicates that social distancing has two effects: it reduces the basic reproduction number of the infection as expected, and shortens the time required for the epidemic to peak. This latter effect is unexpected, as changes in behavior should reduce transmission immediately, which, after a fixed delay – involving manifestation of symptoms and in a fraction of the patients death – should also appear in the daily death toll *D*(*t*). We propose that the increased time between the peak and the time of the social distancing may indicate the presence of subpopulations in which the disease continues to propagate with the initial reproduction number 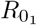. In these populations the transmission is eventually blocked, not by the overall social distancing efforts within the society, but by some other means. As a likely mechanism, we suggest that the local outbreak can reach such a magnitude that it either triggers an intervention or allows the establishment of herd immunity. Prominent examples of such events that collectively expand the duration of the initial growth phase are outbreaks in nursing homes, meat processing plants, warehouses and prisons – which become more likely when the overall social distancing is weak. Furthermore, weak overall social distancing could also fail to protect and segregate these vulnerable subpopulations specifically, thus increase the effective size of such subpopulations.

## 5 Discussion

In this paper we analysed the interdependence of epidemic and mobility data and identified a quantitative relation between parameters of social distancing and key characteristics of the COVID-19 epidemic. Our sample consisted of 9 European countries where suitable data was available at current time. We found that the total death toll does not correlate well with any single parameter such as the timing of the official lock-down or the strictness of social distancing extracted from mobile phone location data. The total death toll, however, could be well estimated by a non-linear combination of three parameters: exponents characterising (i) the initial exponential growth rate (or reproductive number, 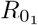 and (ii) the final decay rate of the epidemic, 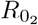, and (iii) the peak death rate 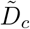 which separates the two stages. The initial growth rate is an intrinsic parameter which may vary somewhat across different countries, but is not affected by control measures or social responses to the epidemic. Based on our data analysis we find that the two remaining parameters, 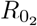 and 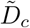 can be related to the timing (*t*_eff_) and strength (*µ*) of social distancing.

Since the estimated daily death toll at the peak increases exponentially with the difference between the effective lock-down time (*t*_eff_) and time of the peak (*t*_*c*_), small changes in *t*_*c*_ − *t*_eff_ can yield substantial differences in the total death toll. The per capita values of 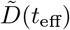 are fairly similar: within the range 0.1-0.16 for 7 of the countries analysed, suggesting that social distancing started at similar stages of the epidemic. The important exception is Germany, where a much smaller value of 0.02 corresponds to roughly a week earlier response. According to the analysis presented here, this explains the substantially lower German death toll, in spite of the relatively moderate social distancing. The slightly higher value of 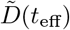/population in Spain (0.23) was compensated by a strict lock-down (lower *µ*).

The higher death toll in Belgium cannot be explained by the proposed set of parameters (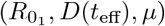) as all three values are close to the average of the sample. This anomaly can be traced to Fig. 5(b), where the deviation of the Belgian data point from the fitted curve indicates that the peak of the epidemic was delayed by approximately 4 days compared to what could be expected based on the proxy measure for social distancing *µ*. While the high Belgian death toll is often attributed to the different methodology of recording COVID-19 related fatalities, including suspected deaths which were not confirmed by lab analysis, such a difference in methodology cannot explain the markedly high time distance between the effective date of social distancing (*t*_eff_) and the epidemic peak (*t*_*c*_); see Fig. 5(b). As we propose that *t*_*c*_ − *t*_eff_ reflects the presence of subpopulations in which the disease can spread unmitigated by social distancing efforts, we suggest that such groups were relatively larger in Belgium than in the other countries within our sample.

While in the case of Sweden the initial growth rate was the slowest and even the timing (*D*(*t*_eff_)) was similar to other countries the weakness of the mobility restriction (i.e. large *µ*) led to high death toll resulting from a strongly delayed peak. We would like to emphasize, that this delay was unexpected at the time of social distancing efforts, and still unaccounted by the typical SIR-based epidemiological models [10].

In the case of the UK the timing of the social distancing (*D*(*t*_eff_)) is similar to other countries, however the mobility restriction appears to be weaker (i.e. higher *µ*) compared to similar countries (Spain, Italy and France) which results in a slower decay of the epidemic.

The phone mobility data is an indirect measure of the social distancing and disease transmission probabilities. However, it can be compared to more traditional measures in the Netherlands, where a study directly determined the number of daily personal interactions before and after the lock-down. Using questionnaires, the study identified a 71% decrease in interactions on average, which is fairly close to the 65% drop estimated from the phone mobility data [14]. While mobile phone tracking data thus can characterize the relatively early stages of social distancing efforts, it may be less useful to detect more subtle efforts like wearing masks or staggered working shifts at later stages of the epidemic.

In conclusion, we demonstrated a quantitative relation between social distancing efforts and statistical parameters describing the current COVID-19 epidemic. We identified an unexpected extension of the exponential growth phase when social distancing efforts are weak, which can substantially increase the death toll of the disease. Future studies are required to extend this analysis to countries with a markedly different socio-economical arrangements.

## Data Availability

The COVID-19 data used in this manuscript is available to the public.

## 6 Supplemental Material

**Table S1:**
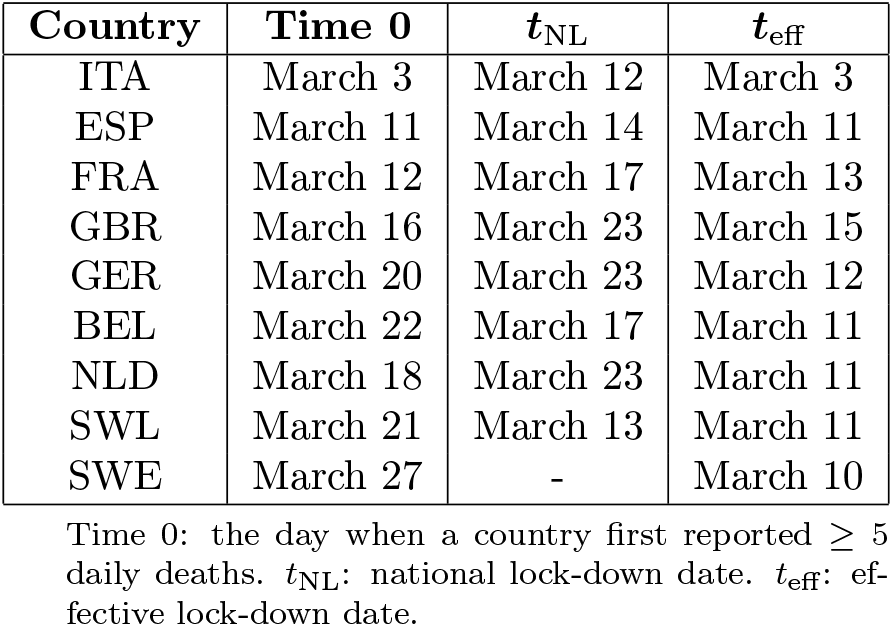
Calendar dates associated with the daily deaths and mobility data.

**Table S2:** Parameters from fitting Equation (1), to the number of daily deaths data during the exponential growth and exponential decay phases. Parameter values correspond to mean ± SE.

| Country | $D_{0_1}$ | $\alpha_1$ | $D_{0_2}$ | $\alpha_2$ |
| --- | --- | --- | --- | --- |
| ITA | $9.934 \pm 0.86$ | $0.25 \pm 0.01$ | $2207.08 \pm 75.03$ | $-0.032 \pm 0.002$ |
| ESP | $9.818 \pm 0.79$ | $0.25 \pm 0.01$ | $2987.16 \pm 321.018$ | $-0.046 \pm 0.002$ |
| FRA | $6.95 \pm 0.50$ | $0.22 \pm 0.01$ | $6970.80 \pm 388.52$ | $-0.058 \pm 0.002$ |
| GBR | $8.59 \pm 0.81$ | $0.22 \pm 0.01$ | $2739.04 \pm 89.85$ | $-0.030 \pm 0.002$ |
| GER | $7.32 \pm 0.47$ | $0.20 \pm 0.01$ | $1089.01 \pm 48.01$ | $-0.046 \pm 0.002$ |
| BEL | $9.97 \pm 0.61$ | $0.20 \pm 0.01$ | $1433.01 \pm 69.99$ | $-0.056 \pm 0.002$ |
| NLD | $7.28 \pm 0.31$ | $0.22 \pm 0.01$ | $851.46 \pm 49.79$ | $-0.052 \pm 0.002$ |
| SWL | $5.25 \pm 0.46$ | $0.19 \pm 0.01$ | $321.90 \pm 25.24$ | $-0.064 \pm 0.002$ |
| SWE | $10.53 \pm 0.61$ | $0.14 \pm 0.01$ | $151.46 \pm 11.16$ | $-0.018 \pm 0.002$ |

**Table S3:** Parameters from fitting Equation (3) to the mobility data over a period of 90 days starting from 13-January-2020. Parameter values correspond to mean ± SE.S

| Country | $M_1$ | $t_1$ | $M_2$ | $t_2$ |
| --- | --- | --- | --- | --- |
| ITA | $116.07 \pm 1.74$ | $-11.00 \pm 1.03$ | $14.45 \pm 2.08$ | $11.01 \pm 1.06$ |
| ESP | $123.87 \pm 1.94$ | $-2.75 \pm 0.72$ | $10.80 \pm 2.77$ | $3.51 \pm 0.69$ |
| FRA | $101.22 \pm 1.24$ | $-1.69 \pm 0.62$ | $16.27 \pm 1.87$ | $4.63 \pm 0.62$ |
| GRB | $115.37 \pm 1.51$ | $-6.84 \pm 1.09$ | $36.64 \pm 2.51$ | $5.20 \pm 1.05$ |
| GER | $112.40 \pm 1.30$ | $-13.44 \pm 1.05$ | $46.14 \pm 1.96$ | $-1.21 \pm 1.12$ |
| BEL | $126.14 \pm 1.74$ | $-13.81 \pm 0.93$ | $39.36 \pm 2.52$ | $-6.37 \pm 0.91$ |
| NLD | $112.78 \pm 1.22$ | $-11.78 \pm 0.87$ | $39.86 \pm 1.78$ | $-1.40 \pm 0.87$ |
| SWL | $108.10 \pm 1.00$ | $-15.25 \pm 0.80$ | $44.72 \pm 1.46$ | $-3.28 \pm 0.87$ |
| SWE | $112.53 \pm 0.95$ | $-21.00 \pm 1.18$ | $70.60 \pm 1.33$ | $-12.06 \pm 1.19$ |

$M_1$ : average mobility level before social distancing. $t_1$ : when the mobility started to decrease due to social distancing. $M_2$ : average mobility after the social distancing. $t_2$ : end of the transition period to social distancing.

